# The pleiotropic contribution of genes in dopaminergic and serotonergic pathways to addiction, aggression, and related behavioural traits

**DOI:** 10.1101/2023.05.03.23289424

**Authors:** Ester Antón-Galindo, Judit Cabana-Domínguez, Bàrbara Torrico, Roser Corominas, Bru Cormand, Noèlia Fernàndez-Castillo

## Abstract

Co-occurrence of substance use disorders (SUD) and aggressive behaviour in the same individual has been frequently described. As dopamine (DA) and serotonin (5-HT) are key neurotransmitters for both phenotypes, we explored the genetic contribution of these pathways to SUD, aggressive behaviour and related behavioural traits. Here, we tested the association of 275 dopaminergic genes and 176 serotonergic genes with these phenotypes by performing gene-based, gene-set and transcriptome-wide association studies (TWAS) in 11 genome-wide association studies (GWAS) datasets on SUD (alcohol, cocaine, cannabis, opioids and a multivariate analysis of three drugs of abuse), aggressive behaviour (disruptive behaviour and antisocial behaviour) and related behaviours (irritability, neuroticism, risk taking and anxiety). At the gene-wide level, 68 DA and 27 5-HT genes were found to be associated with at least one GWAS on SUD or related behaviour. Among them, six genes had a pleiotropic effect, being associated with at least three phenotypes: *ADH1C, ARNTL, CHRNA3, HPRT1, HTR1B* and *DRD2*, the latter with five. Additionally, we found nominal associations between the DA gene sets and antisocial behaviour, opioid use disorder, SUD, irritability and neuroticism, and between the 5-HT-core gene set and neuroticism. Gene expression correlates in brain were also found for 19 genes, highlighting the association for *CHRNA3* and *CELSR3* with OUD, SUD and irritability and *CELSR3* also with neuroticism. Our study shows a pleiotropic contribution of dopaminergic and serotonergic genes to addiction, aggression and related behaviours, highlighting a special role for DA genes, which could explain, in part, the co-occurrence of these phenotypes.

## INTRODUCTION

Addiction is a complex chronic disorder that impacts millions of people around the world (Substance Abuse and Mental Health Services Administration, 2022). Clinically, addiction is now encompassed by the term substance use disorders (SUD) and is characterized in DSM-5 by a core set of behavioural features that can be grouped into impaired control of substance use, impaired social behaviour and risky substance use (American Psychiatric Association, 2013). Recent research has pointed to an association between SUD and aggressive behaviour, as these conditions are frequently co-occurring (Ghossoub et al., 2019; von der Pahlen et al., 2008). While aggressive behaviour confers some evolutionary advantages, its expression in the wrong context can lead to injury, harm and serious social problems (Anholt and MacKay, 2012; de Almeida et al., 2015).

Comorbidity between these two conditions is complex. On the one hand, substance use can directly influence aggressive behaviour (through disinhibitory and acute effects) or indirectly (through exposure to violence, personality traits and social context) (Hoaken and Stewart, 2003). An increased risk for abnormal or pathological aggression has been described in individuals diagnosed with psychiatric disorders, including substance use disorders (Ghossoub et al., 2019; Hoaken and Stewart, 2003; Kalk et al., 2022). More precisely, several drugs of abuse, such as alcohol, cocaine, amphetamines and cannabis, have been related to a higher risk for aggressive behaviour (Boles and Miotto, 2003; Hoaken and Stewart, 2003). On the other hand, it is also possible that aggressive behaviour increases risk for substance use or addiction, and underlying biological mechanisms may contribute to both disorders. Indeed, greater polygenic risk for aggression was found to increase risk for substance use offending and SUD diagnosis (Elam et al., 2021). Also, a clinical study found that risk for SUD in individuals with intermittent explosive disorder was much higher than in controls, and that this aggressive behaviour was identified before SUD diagnosis in 80% of cases (Coccaro et al., 2016).

Transdiagnostic behavioural traits, such as irritability, neuroticism, anxiety or risk-taking behaviour, have been frequently described in individuals with SUD or with aggressive behaviour. In adults, irritability is regarded as a feature of substance use, disruptive, antisocial and conduct disorders among others, and can be a precursor to aggression in some cases (Beauchaine and Tackett, 2020; Bell et al., 2021; Toohey and DiGiuseppe, 2017). Neuroticism is a robust personality trait characterized by emotional instability and is often associated with a higher risk for developing psychiatric disorders (Lahey, 2009). Anxiety has been related to conduct problems and reactive relational aggression (Cunningham and Ollendick, 2010; Marsee et al., 2008), and previous research has reported an association between SUD and independent anxiety disorders (Grant et al., 2004). Finally, risk-taking behaviour has been closely linked to SUD and aggressive behaviour and violence, involving a preference for moderate or high short-term rewards with the potential for a great loss, which is perceived as exciting (Leather, 2009). Although these behaviours have been reported in individuals with SUD or aggressive behaviour, the common genetic and neurobiological factors explaining this co-occurrence are not yet fully understood.

Over the years, research efforts have been made to characterize the neurobiological and psychological underpinnings of SUD and aggression. It is clear that both addiction and aggressive behaviour are multifactorial disorders, where genetic variation and environmental factors play a role in their development (Agrawal et al., 2012; Hancock et al., 2018; Popescu et al., 2021; Veroude et al., 2016; Vink, 2016). Both SUD and aggression cause rewarding effects, and several studies have investigated potential shared neurobiological mechanisms between them, suggesting the reward pathway as a common underlying mechanism (Chester and DeWall, 2016; Golden and Shaham, 2018). Indeed, dopamine and serotonin are key neurotransmitters in reward and have been widely related to SUD and aggressive behaviour (Couppis and Kennedy, 2008; Golden et al., 2019).

The critical role of DA and 5-HT in addiction processes has been extensively demonstrated over more than 40 years (Koob and Volkow, 2016; Müller and Homberg, 2015; Solinas et al., 2019). DA circuits are key modulators of behaviours associated with SUD via different mechanisms, and nearly all drugs used by humans acutely increase DA signalling within the striatum (Poisson et al., 2021; Wise and Jordan, 2021; Wise and Robble, 2020). Additionally, DA signalling is involved in several processes that contribute to the development of addiction, such as reward, learning and motivation (Wise and Jordan, 2021; Wise and Robble, 2020). In aggressive behaviour, DA neurotransmission plays an important role in reward related to aggression, and neurons of medial hypothalamic and mesolimbic circuits modulate this behaviour (Yamaguchi and Lin, 2018). Evidence from behavioural and neuroimaging studies has pointed to a neural imbalance in the reward pathway being involved in retaliatory aggression (Chester and DeWall, 2016), and research on animal models has described a major contribution of the dopaminergic reward circuitry to appetitive aggression and relapse to aggression seeking (Couppis and Kennedy, 2008; Golden et al., 2019; Golden and Shaham, 2018).

In addition, 5-HT is a key neurotransmitter with a major role in aggressive behaviour largely confirmed by decades of research (Coccaro et al., 2015). 5-HT modulates the activity of specific brain areas involved in the control of limbic response, and individuals with increased aggressive behaviour have impaired serotonergic functioning in these regions (Coccaro et al., 2015). Also, the essential involvement of the 5-HT system in both the establishment of drug use-associated behaviours and the transition and maintenance of addiction has been largely studied (Kirby et al., 2011; Müller et al., 2007; Müller and Homberg, 2015). 5-HT is involved in synaptic plasticity, hedonic tone, motivational and reinforcement processes, learning and memory, all of which are critical processes in the development of addiction (Kirby et al., 2011; Müller et al., 2007; Müller and Homberg, 2015).

In the present study, we aim to comprehensively assess the genetic contribution of the dopaminergic and serotonergic systems to SUD and aggressive behaviour, as well as to other related behavioural traits such as irritability, neuroticism, anxiety and risk-taking behaviour. These phenotypes co-occur frequently in individuals, and our analyses may contribute to better understand of the genetic basis of the comorbidities.

## EXPERIMENTAL PROCEDURES

### DA and 5-HT gene selection

To comprehensively explore all genes involved in dopaminergic and serotonergic pathways, we used four gene sets previously described by us. Two core gene sets and two wide gene sets were elaborated for dopamine (DA) and serotonin (5-HT) as reported by Cabana-Domínguez et al. (Cabana-Domínguez et al., 2022). The two core gene sets, DA-core with 12 genes and 5-HT-core with 23 genes, were obtained through manual curation and contain the well-known dopaminergic and serotonergic genes, including neurotransmitter receptors, transporters, and enzymes involved in their anabolism or catabolism. The two wide gene sets were defined using GO (Gene Ontology Consortium, http://geneontology.org/) and KEGG (https://www.genome.jp/kegg/pathway.html) datasets, obtaining a DA-wide set of 275 genes and a 5-HT-wide set of 176 genes (Supplementary Table 1). The intersection of these lists includes 57 genes that participate in both dopaminergic and serotonergic pathways, one of which belongs to the core sets.

Since only some of the summary statistics used included the X chromosome (anxiety, irritability and neuroticism), 14 genes located in this chromosome could not be tested in most of the datasets: 9 from DA gene sets (*ATP7A, AGTR2, FLNA, GPR50, GRIA3, HPRT1, MAOA, MAOB* and *PPP2R3B*) and 8 from the 5-HT gene sets (*ATP7A, ARAF, ASMT, CACNA1F, GPM6B, HTR2C, MAOA* and *MAOB*), being three of them present in both.

### Data used from GWAS of addiction, aggressive behaviour and related traits

In this study, we used publicly available data from different studies of SUD, aggressive behaviours and related traits. We used a total of 19 summary statistics of genome-wide association studies (GWAS) performed in individuals with European ancestry, including 8 datasets of SUD, three of aggressive behaviour and 8 of related behavioural traits. Data were either downloaded from the Psychiatric Genomics Consortium (PGC) (https://pgc.unc.edu/for-researchers/download-results/), the UKBiobank (https://www.ukbiobank.ac.uk/), the iPSYCH (https://ipsych.dk/en/research/downloads) or the BroadABC (http://broadabc.ctglab.nl/summary_statistics) web pages, or shared by the authors of the GWAS (details in Supplementary Table 2).

### Selection of summary statistics based on heritability analyses and variant filtering

The SNP heritability of the 19 GWAS mentioned above (Supplementary Table 2) was estimated using linkage disequilibrium score regression (LDSC) (https://github.com/bulik/ldsc) (Bulik-Sullivan et al., 2015). In the case of *cocaine dependence, alcohol dependence, cannabis dependence, cannabis use disorder, opioids dependence, opioids use disorder, ever addicted* phenotype, and *anxiety*, heritability was reported on the liability scale considering the sample and population prevalence of each of them. For the other GWAS, a liability scale could not be used due to the absence of a population prevalence estimate or the use of a continuous scale to define the traits.

A total of 7 GWAS were discarded with a SNP-based heritability estimates h^2^_SNP_ <0.05, indicating a low genetic contribution. In the case of *opioids addiction*, both summary statistics of *opioids dependence* and *opioids use disorder* showed a heritability higher than 5%, but *opioids use disorder* summary statistics was selected for subsequent analyses due to a higher number of individuals included in this study (Supplementary Table 2).

In total, 11 summary statistics from GWAS were selected for subsequent analyses, including 5 studies on SUD (*alcohol dependence* (Walters et al., 2018), *cannabis use disorder* (CUD) (Johnson et al., 2020), *cocaine dependence* (Cabana-Domínguez et al., 2019), *opioids use disorder* (OUD) (Deak et al., 2022) and a multivariate analysis of three *substance use disorders* (SUD) (Schoeler et al., 2022)), two on aggressive behaviour (*antisocial behaviour* (AB) (Tielbeek et al., 2017) and *disruptive behaviour disorders comorbid with ADHD* (ADHD-DBD) (Demontis et al., 2021)) and four on related behavioural traits (*risk taking behaviour, irritability, anxiety* (Meier et al., 2019), and *neuroticism*).

Genetic variants from most of the summary statistics used were filtered out by MAF ≤ 0.01 and info-score for imputation quality ≤ 0.8. There were three exceptions in which variants with a lower imputation quality could not be filtered out because the specific info-score values were missing: antisocial behaviour (info-score>0. 6), risk taking (info-score>0.4), and SUD.

### Gene-based and gene-set analyses

The contribution of common variants in the DA/5-HT-related genes to SUD, aggressive behaviour or related behavioural traits was assessed through gene-based and gene-set analyses using the 11 GWAS summary statistics selected.

Gene-based association studies were performed on MAGMA v1.10 (de Leeuw et al., 2015) using the SNP-wise mean model, with the test statistic being the sum of -log(SNP p value) for SNPs located within the transcribed region (defined on NCBI 37.3 gene definitions). The analysis was performed without window around the gene using the 1000 Genomes Project Phase 3 (European data only) as a reference panel (Auton et al., 2015). False Discovery Rate (FDR) was used to correct for multiple testing (5% FDR).

Competitive gene-set analyses were performed for the four sets of genes (DA-core, DA-wide, 5-HT-core and 5-HT-wide) using MAGMA to assess their association with the studied phenotypes. Multiple-testing Bonferroni correction was applied considering 44 gene-set tests (p<0.0011).

### Effect on brain volumes

To investigate the effect on brain volumes, summary statistics of GWAS meta-analysis in individuals of European ancestry of 7 subcortical volumes (amygdala, caudate nucleus, hippocampus, nucleus accumbens, pallidum, putamen and thalamus) and intra-cranial volume (13,171 individuals) (Hibar et al., 2015), and cortical thickness and brain surface area (23,909 individuals) (Grasby et al., 2020) were downloaded from the ENGIMA web page (https://enigma.ini.usc.edu/). Gene-based and gene-set analyses were performed in MAGMA for each volume as previously described.

### Gene expression correlates in each phenotype

We considered all the SNPs located in each DA and 5-HT gene to infer whether the genetically-predicted expression of each DA and 5-HT gene correlates with the 11 phenotypes of this study. These analyses were carried out on MetaXcan (S-PrediXcan (Barbeira et al., 2021, 2018; Gamazon et al., 2015) and S-MultiXcan (Barbeira Id et al., 2019)) using the summary statistics of each disorder or trait. Prediction elastic-net models were downloaded from PredictDB (https://predictdb.org/post/2021/07/21/gtex-v8-models-on-eqtl-and-sqtl/), which were constructed considering SNPs located within 1 Mb upstream of the transcription start site and 1 Mb downstream of the transcription end site of each gene and were trained with RNA-Seq data of 13 GTEx (release V8) brain regions: amygdala, anterior cingulate cortex BA24, caudate, cerebellar hemisphere, cerebellum, cortex, frontal cortex BA9, hippocampus, hypothalamus, nucleus accumbens, putamen, spinal cord and substancia nigra. S-PrediXcan was used to analyse the genetically determined expression of genes in each of the 13 brain tissues for each of the 11 phenotypes. Then, the information across tissues was combined for each phenotype using a multivariate regression with S-MultiXcan, and a multiple-testing FDR correction (5% FDR) was applied.

## RESULTS

### DA and 5-HT genes are associated with addiction, aggression and related behaviours

We conducted a comprehensive study to explore the contribution of genes involved in dopaminergic and serotonergic pathways to addiction, aggression and related behaviours. Through gene-based analyses we investigated the association of a total of 275 genes in the DA-wide set and 176 genes in the 5-HT-wide set (57 of them included in both pathways) (Supplementary Table 1) with SUD (*alcohol dependence, cannabis use disorder* (CUD), *cocaine dependence, opioids use disorder* (OUD) and a multivariate analysis of three substance use disorders (SUD)), aggressive behaviour (*antisocial behaviour* (AB) and *attention-deficit and hyperactivity disorder comorbid with disruptive behaviour* (ADHD-DBD)) and four on related behavioural traits (*risk taking behaviour, irritability, anxiety* and *neuroticism*).

At the gene-wide level, several genes from both the DA-wide and 5-HT-wide sets were found to be significantly associated (overcoming a multiple testing correction of FDR 5%) with 7 of the analysed phenotypes (Figure 1). However, most of the SUD or aggression GWAS lacked power, as shown by the limited number of associated genes in total, and we could not identify associated genes in the DA and 5-HT gene sets (Figure 1A).

**Figure 1.**
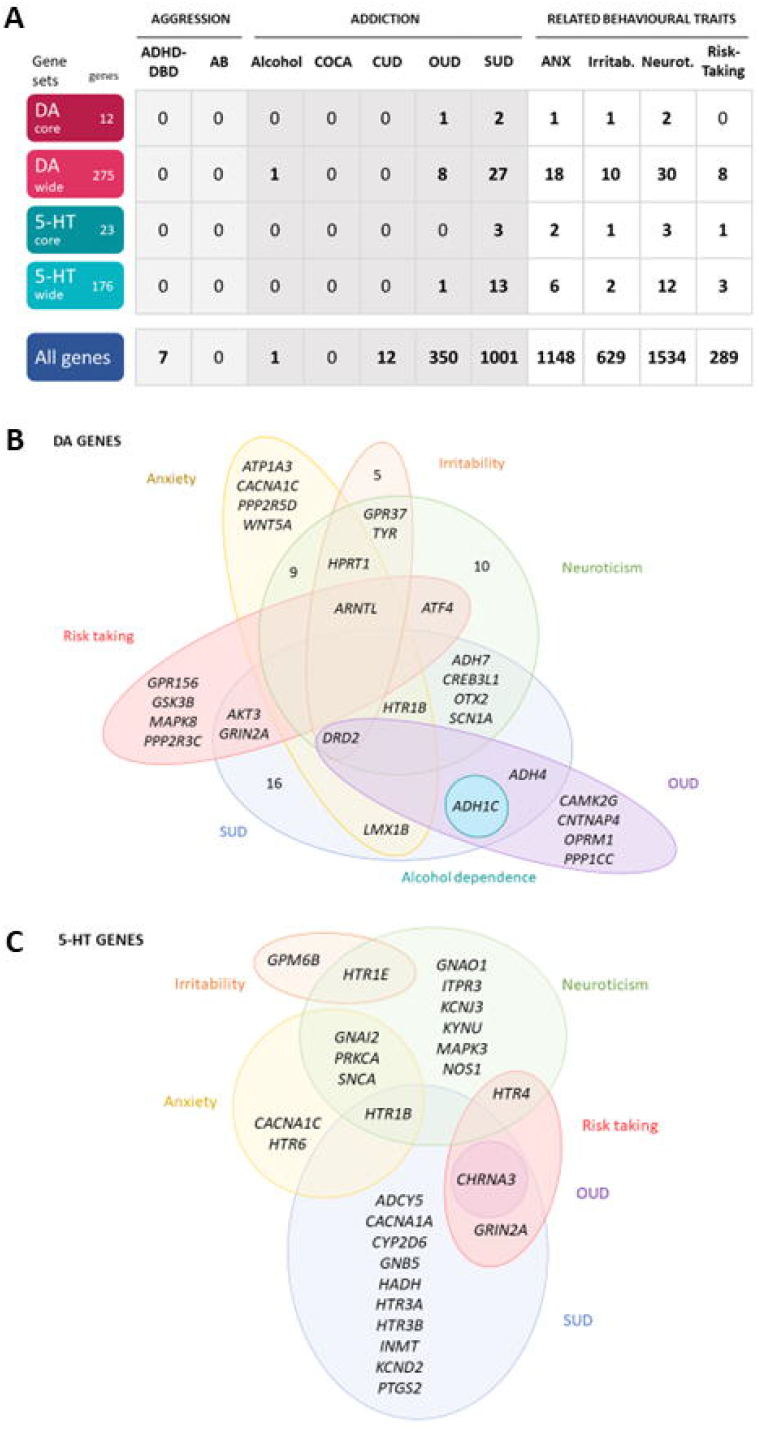
Dopaminergic and serotonergic genes associated with the studied disorders or traits. **A)** Number of significantly associated genes in the gene-based analyses of 11 disorders or traits. 5-HT, serotonin; DA, dopamine. In light grey, aggressive behaviours; in dark grey, addiction disorders; in white, related behavioural traits. **B and C)** Venn diagrams of the significantly associated **B)** dopaminergic (DA) and **C)** serotoninergic (5-HT) genes across the studied disorders or traits. ADHD-DBD, attention-deficit and hyperactivity disorder comorbid with disruptive behaviour; AB, antisocial behaviour; Alcohol, alcohol dependence; ANX, anxiety; COCA, cocaine dependence; CUD, cannabis use disorder; Irritab., irritability; Neurot., neuroticism; OUD, opioids use disorder; SUD, substance use disorder; Risk-Taking; risk-taking behaviour. All significant genes overcome a multiple-testing correction of 5% False Discovery Rate, FDR.

Among the DA-wide gene set, one gene was found to be significantly associated with alcohol dependence, 8 with OUD, 27 with SUD, 18 with anxiety, 10 with irritability, 30 with neuroticism and 8 with risk taking (Figure 1A and B, and Supplementary Table 4). Interestingly, five DA genes were found associated with at least three phenotypes, showing a pleiotropic effect: *DRD2, ARNTL, ADH1C, HPRT1* and *HTR1B* (Figure 1A, Supplementary Table 4). In particular, *HPRT1* was associated with the only three phenotypes in which it could be tested: anxiety (p=4.34E-04), irritability (p=2.05E-06) and neuroticisim (p=5.70E-08), since this gene is located in the X chromosome. Three DA core genes were found to be significantly associated with at least one disorder: *DRD2* is associated with OUD (p=6.25E-08), SUD (p=7.74E-13), anxiety (p=3.10E-05), irritability (p=3.41E-05) and neuroticism (p=6.12E-13); *DBH* is associated with SUD (p=3.20E-04); and *DRD3* is associated with neuroticism (p=3.38E-03) (Supplementary Table 4).

Among the 5-HT-wide genes, one gene was found to be significantly associated with OUD, 13 with SUD, 6 with anxiety, 2 with irritability, 12 with neuroticism and 3 with risk taking (Figure 1A and C, and Supplementary Table 5), some of them also present among the DA genes. Interestingly, two 5-HT genes, *HTR1B* and *CHRNA3*, were found significantly associated with three phenotypes (Figure 1A and C and Supplementary Table 5). Six 5-HT-core genes were found to be significantly associated with at least one disorder: *HTR1B* with SUD (p=3.43E-03), anxiety (p=4.34E-04) and neuroticism (p=9.63E-04), *HTR1E* with irritability (p=1.82E-04) and neuroticism (p=5.89E-05), *HTR4* with neuroticism (p=5.21E-03) and risk taking (p=1.15E-03), *HTR6* with anxiety (p=1.71E-03), and *HTR3A* and *HTR3B* with SUD (p=1.17E-03 and p=1.45E-03, respectively) (Supplementary Table 5).

Finally, we performed gene-set analyses to assess the contribution of the four gene sets to each of the 11 phenotypes studied (Figure 2). Interestingly, we found that the DA-wide gene set was nominally associated with antisocial behaviour (p=0.022), OUD (p=0.020) and SUD (p = 0.017), and that the DA core gene set was nominally associated with SUD (p= 0.021), irritability (p=0.006) and neuroticism (p=0.041). On the other hand, the 5-HT core gene set showed exclusively a nominal association with neuroticism (p=0.007) and the 5-HT-wide gene set did not show association with any trait. None of these associations overcame Bonferroni correction.

**Figure 2.**
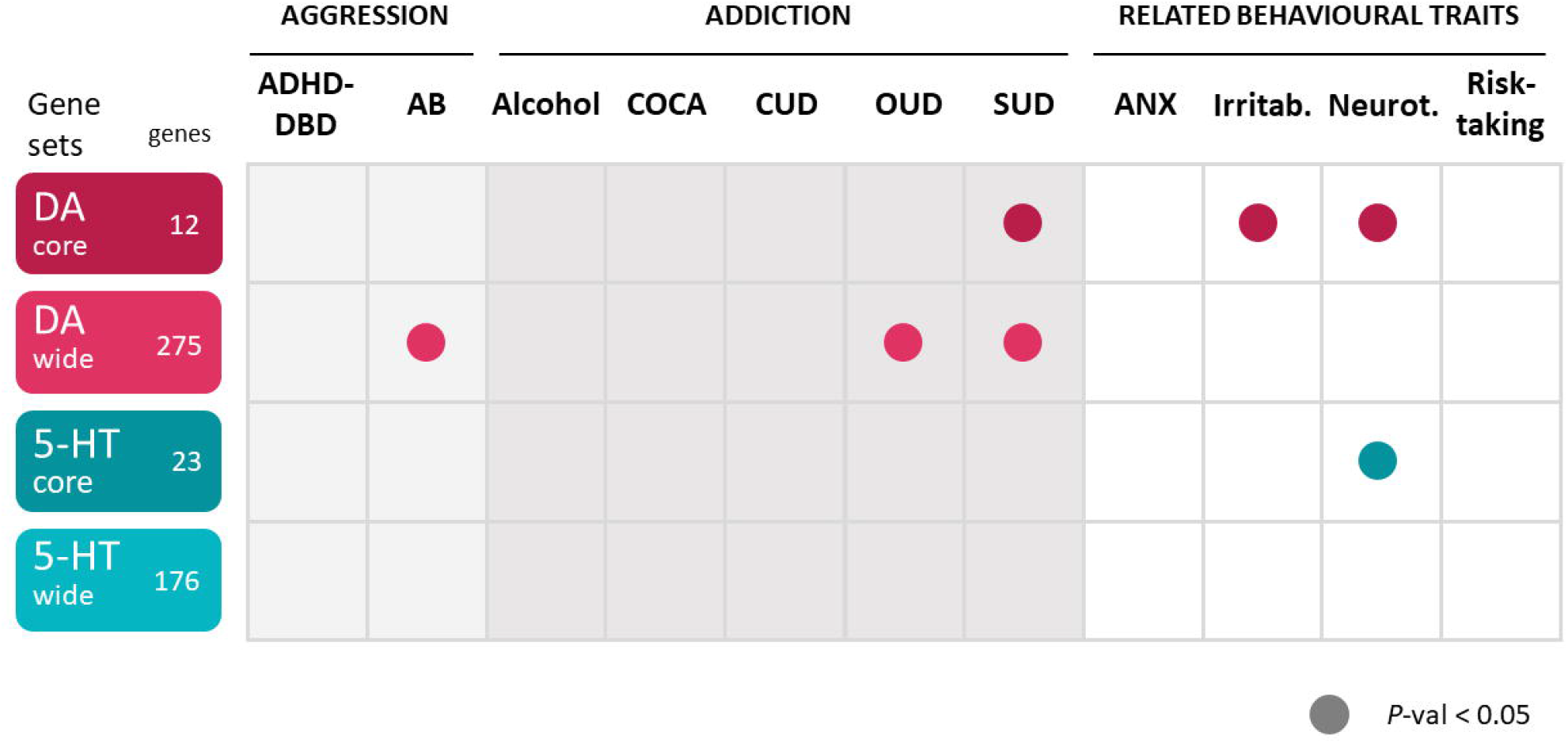
Significant associations of the dopamine and serotonin gene sets with 11 disorders or traits. 5-HT, serotonin; DA, dopamine. ADHD-DBD, attention-deficit and hyperactivity disorder comorbid with disruptive behaviour; AB, antisocial behaviour; Alcohol, alcohol dependence; ANX, anxiety; COCA, cocaine dependence; CUD, cannabis use disorder; Irritab., irritability; Neurot., neuroticism; OUD, opioids use disorder; SUD, substance use disorder; Risk-Taking; risk-taking behaviour. P-val, p-value.

### DA and 5-HT genes associated with volumetric brain changes

We also investigated whether the DA and 5-HT genes were associated with volumetric brain changes by performing a gene-based analysis using data from the ENIGMA consortium. Only the cortical thickness and surface area measures showed significant associations with four DA genes and one 5-HT gene, and no associations were found for the other measures in subcortical regions, probably due to lack of power (Supplementary Figure 1). *AKT3* and *GPR21*, belonging to the DA-wide gene set, and *DDC*, belonging to both DA core and 5-HT core gene sets, were significantly associated with alterations in surface area (p=4.87E-05, p=2.27E-05 and p=2.27E-04, respectively). In addition, *ATP1A3*, from the DA-wide set, was associated with alterations in cortical thickness (p=5.94E-05). Finally, we performed gene-set analyses and found only a nominal association of the DA-wide gene set with cortical thickness (p=0.015). Interestingly, one of these three genes, *AKT3*, which was associated with alterations in surface area and cortical thickness, was also associated with SUD and risk taking (Supplementary Table 4).

### Gene expression correlates in brain regions with addiction, aggression and related behaviours

Finally, we explored whether the predicted expression of DA and 5-HT genes correlated with the 11 phenotypes analysed previously using S-PrediXcan and S-MultiXcan analyses. Interestingly, the expression of 14 DA, 4 5-HT genes and one gene belonging to both DA and 5-HT gene sets was found to be significantly associated with the phenotype for OUD, SUD, anxiety, irritability or neuroticism (Table 1), and almost all of them were also significantly associated with at least one phenotype in the gene-based analysis. Among them, 7 genes were found to be associated with two traits, the 5-HT gene *CHRNA3* with three traits (OUD, SUD and irritability), and the DA gene *CELSR3* with four traits (OUD, SUD, irritability and neuroticism). We did not find any gene significantly associated with aggressive behaviour (antisocial behaviour or ADHD-DBD), probably due to the lack of statistical power, and no DA and 5-HT genes were found significantly associated with CUD and risk-taking (Supplementary Table 6).

**Table 1.**
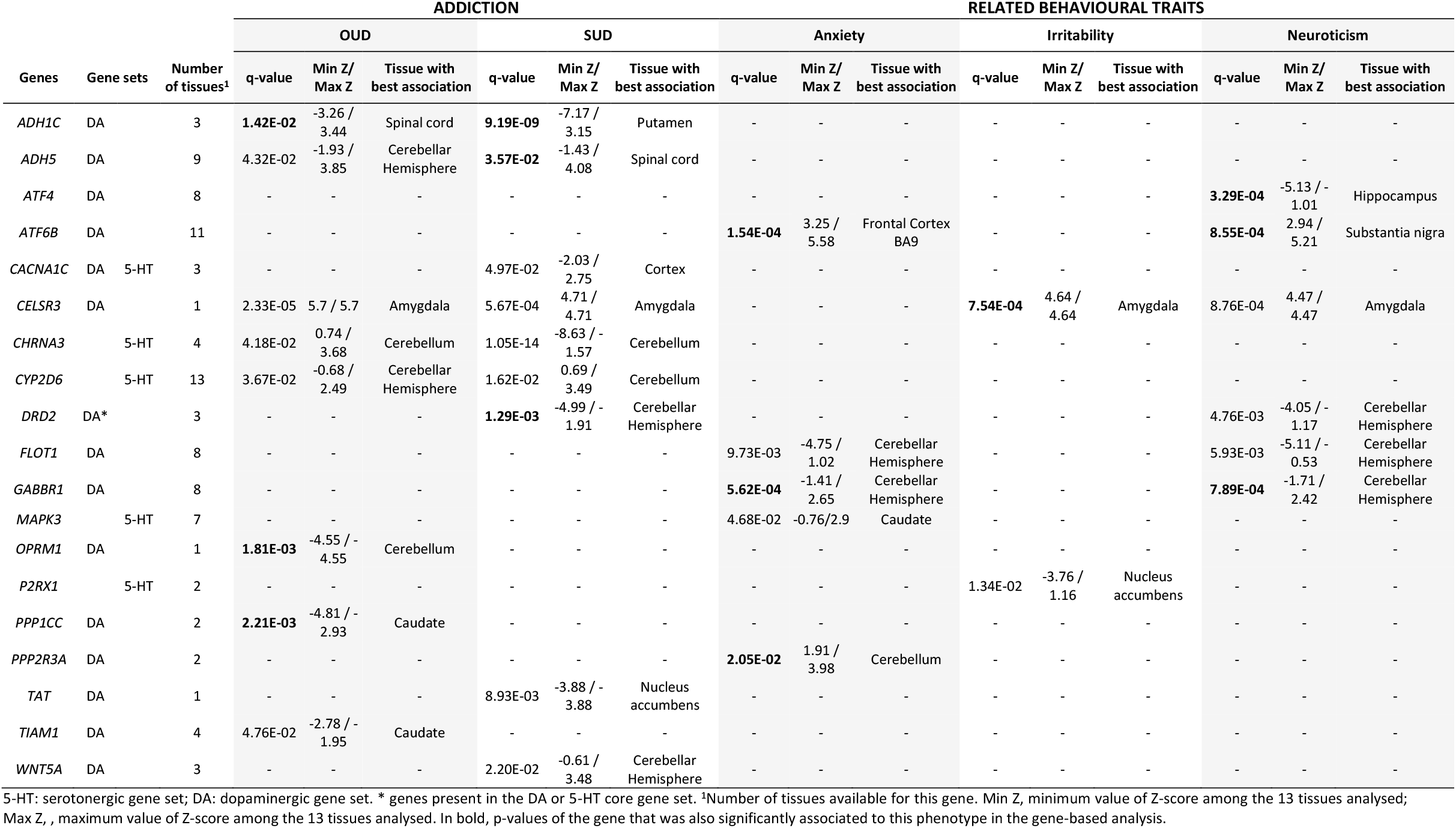
Genes from the dopaminergic and serotoninergic gene sets significantly associated to at least one disorder or trait in the TWAS analysis.

## DISCUSSION

Dopaminergic and serotonergic neurotransmission have been widely implicated in both SUD and aggressive behaviour, as described above. Decades of research on animal models and candidate gene association studies have pointed to genes that encode proteins involved in the dopaminergic and serotonergic systems as some of the main genetic contributors to these phenotypes (Anholt and MacKay, 2012; Fernàndez-Castillo and Cormand, 2016; Le Foll et al., 2009; Palmer and De Wit, 2012; Rosell and Siever, 2015; Veroude et al., 2016; Zhang-James et al., 2019). However, the vast majority of these association studies, which investigated genetic variants in the core DA and 5-HT genes, were performed in small samples, and the lack of power can explain the contradictory findings or false positive associations (Fernàndez-Castillo et al., 2021; Fernàndez-Castillo and Cormand, 2016; Gelernter and Polimanti, 2021).

This is the first systematic genetic study of the DA and 5-HT pathways in different SUDs, aggressive behaviours and related behavioural traits. Here, we assessed the contribution of common genetic variation of a comprehensive list of genes involved in DA and 5-HT neurotransmission (275 and 176 genes, respectively) to these disorders and behavioural traits. Our results show a genetic contribution of both pathways to these phenotypes, suggesting a major role for dopamine genes, as pinpointed by the gene-set analyses. We found several genes from both pathways, mostly DA but also 5-HT, associated with the studied phenotypes. Remarkably, 6 of them showed pleiotropic effects, with significant associations with SUD and other related behaviours: *ARNTL, ADH1C, CHRNA3, DRD2, HPRT1* and *HTR1B*. Altered expression was predicted for some phenotypes for three of them: *ADH1C, CHRNA3* and *DRD2*.

Dopamine gene sets were associated with antisocial behaviour, OUD and SUD (DA-wide set), and irritability, neuroticism and SUD (DA-core set), highlighting a role for dopaminergic genes. These findings are in line with previous results in other psychiatric disorders obtained by our group with the same gene sets, in which we found association of dopaminergic gene sets with ADHD, autism, bipolar disorder, major depressive disorder, Tourette’s syndrome, schizophrenia and a cross-disorder meta-analysis (Cabana-Domínguez et al., 2022). In both studies, most of the genes found associated are not core genes, *DRD2* being the only exception. These results show the relevance of inspecting genes involved in dopamine or serotonin neurotransmission that are not only core genes (receptors, enzymes and transporters). It is well known that the DA system is involved in modulating neural networks of social behaviour (Kopec et al., 2019; Modi and Sahin, 2019). Indeed, the mesolimbic DA system, involved in reward, is hypothesised to mediate the reinforcing effects of social interactions and drive social motivation in both rodents and humans (Gunaydin et al., 2014; Modi and Sahin, 2019) and is also a key factor during the onset of addiction (Pascoli et al., 2015). Also, dopaminergic antagonists are the most commonly used treatment for suppressing human aggression in psychotic patients (Yamaguchi and Lin, 2018). Moreover, irritability is a core feature of mood disorders, and evidence relates this trait with aberrant striatal responses to DA and low striatal DA levels (Beauchaine and Tackett, 2020; Bell et al., 2021). Finally, several polymorphisms in the *COMT* gene, encoding the enzyme that inactivates catecholamine neurotransmitters, including DA, were previously related to neuroticism and aggressive behaviour (Fernàndez-Castillo and Cormand, 2016; Servaas et al., 2017).

On the other hand, our gene-set analyses revealed only an association related to the 5-HT genes, specifically between the 5-HT-core gene set and neuroticism. This is in line with previous work showing that thalamic 5-HT transporter binding potentials were associated with neuroticism in both males and females, although with opposite directions (Tuominen et al., 2017). Also, polymorphisms in the serotonin transporter were associated with alterations in subnetworks related to cognitive control in women with variable neuroticism scores (Servaas et al., 2017).

When exploring the specific DA and 5-HT genes significantly associated with any of the considered disorders or traits, we found that several of these monoaminergic-related genes were associated with alcohol dependence, OUD and SUD, as well as with the four behavioural traits assessed. Unfortunately, in the ADHD-DBD and the antisocial behaviour GWAS summary statistics almost no gene-wide association was observed. This prevented us from identifying specific shared candidate genes and disentangling the genetic relationship between addiction and aggression.

Remarkably, the most pleiotropic effect was identified for the *DRD2* gene of the dopaminergic core set, encoding the DA receptor D2, which was associated with OUD, SUD, anxiety, irritability and neuroticism. Indeed, in the GWAS of SUD, the top genetic variant operating through the common liability was located on *DRD2* (Schoeler et al., 2022), and *DRD2* showed also a pleiotropic effect in psychiatric disorders being the only gene from the dopaminergic core set associated in the cross-disorder meta-analysis (Cabana-Domínguez et al., 2022). DRD2 has been widely demonstrated to modulate the effects of several drugs of abuse in animal models, especially opiates, alcohol and cocaine (Le Foll et al., 2009). Also, systemic injections of Drd2 antagonists were effective reversing the aggressive phenotype in highly aggressive mice (Fragoso et al., 2016). Finally, a recent study identified a polymorphism in the *DRD2* gene as related to both anxiety and neuroticism scores in patients with polysubstance use disorder (Suchanecka et al., 2020). Another gene showing pleiotropic effects is *CHRNA3*, associated with OUD, SUD and risk-taking behaviour. This gene encodes an acetylcholine receptor and has been widely associated with nicotine dependence and other SUD (Gelernter and Polimanti, 2021). The expression of both *CHRNA3* and *DRD2* was predicted to be altered in multiple brain areas in our TWAS analyses, being the cerebellum and the cerebellar hemisphere the tissues with the best association, respectively (Table 1). Another core gene, *HTR1B*, encoding a serotonin receptor, was associated with SUD, anxiety, and neuroticism. This gene is involved in both DA and 5-HT pathways and associations of genetic variants in it were associated with different SUD but also with aggressive behaviour, anger and hostility (Cao et al., 2013; Fernàndez-Castillo and Cormand, 2016). In the case of *HPRT1*, a gene located on chromosome X, was associated with the three phenotypes that included this chromosome in the GWAS summary statistics, so it could not be assessed in the majority of the summary statistics. *HPRT1* encodes an important enzyme involved in purine nucleotide exchange that is highly expressed in the central nervous system. In our gene-based analyses, *HPRT1* was associated with anxiety, irritability and neuroticism. Interestingly, alterations in HPRT1 function lead to the Lesch-Nyhan syndrome (LNS), a disorder that presents with nervous systems impairments. In previous studies, a dopamine imbalance was shown for LNS models and post-mortem brain from patients, with up to 70-90% decrease of DA levels (Lloyd et al., 1981; Nyhan, 2000; Sculley et al., 1992). *ARNTL*, encoding a transcription factor, is a clock gene essential for the circadian rhythm that has been previously related to psychiatric disorders (Charrier et al., 2017; Kim et al., 2015). In our gene-based analyses, *ARNTL* was associated with the four behavioural traits analysed: anxiety, irritability, neuroticism and risk-taking. Finally, *ADH1C*, encoding alcohol deshydrogenase 1C, is found to be associated with the three addiction disorders alcohol dependence, OUD and SUD. This can be explained by the inclusion of an alcohol use disorder sample in the SUD summary statistics and the probable co-occurrence of alcohol use disorder in the individuals included in the OUD sample. Also, the expression of *ADH1C* was predicted to be altered in multiple brain areas in the OUD and SUD TWAS analyses.

Our results point to a genetic contribution of both DA and 5-HT systems to SUD and related behavioural traits, highlighting a role for DA neurotransmission, which could explain in part their co-occurrence. More genetic studies on aggressive behaviour should be assessed in the future to confirm the contribution of dopaminergic and serotonergic genes to these phenotypes and to better understand the pleiotropic effects of these genes on addiction and other behaviours.

## Data Availability

All data produced in the present study are available upon reasonable request to the authors.

## ACKNOWLEDGEMENTS

We thank all the authors that kindly shared their GWAS data for performing our analyses.

## CONTRIBUTORS

N.F-C. conceived the study and obtained funding, coordinated and administered the project and supervised the analyses, and contributed to writing the first draft of the manuscript. E.A-G. designed and conducted the bioinformatic analyses and wrote the first draft of the manuscript. J.C-D contributed to the study design and bioinformatic analyses. B.C. obtained funding, and together with R.C, supervised and reviewed the manuscript preparation. All authors contributed and have approved the final manuscript.

