## Supplementary Material for "The pleiotropic contribution of genes in dopaminergic and serotonergic pathways to addiction, aggression, and related behavioural traits"

1. **SUPPLEMENTARY TABLES 2 TO 6**

2. **SUPPLEMENTARY FIGURE 1**

3. **REFERENCES**

### 1. SUPPLEMENTARY TABLES

**Supplementary Table 2.** Summary statistics used for the heritability analyses.

| | Trait or disorder | Paper | Source | Individuals |
|---|---|---|---|---|
| ADDICTION | Alcohol dependence | (Walters et al., 2018) | PGC | 11,569 cases + 34,999 controls |
| | Cannabis dependence | (Sherva et al., 2016) | Authors | 1,822 cases + 5,039 controls |
| | Cannabis use disorder | (Johnson et al., 2020) | PGC | 17,068 cases + 357,219 controls |
| | Cocaine dependence | (Cabana-Domínguez et al., 2019) | Authors | 2,085 cases + 4,293 controls |
| | Ever addicted to any substance or behaviour | - | UKBiobank | 7,000 cases + 11,000 controls |
| | Opioids dependence | (Polimanti et al., 2020) | PGC | 3,272 cases + 25,437 controls |
| | Opioids use disorder | (Deak et al., 2022) | Authors | 15,251 cases. 554,186 total individuals |
| | Substance use disorder | (Schoeler et al., 2022) | Authors | Ntot= 187,062, Alcohol use disorder (n=28,757), cannabis use disorder (n=358,534), nicotine dependence (n=244,890), frequency of cigarette (n=245,876), alcohol use (n=513,208), cannabis use (n=24,798) |
| AGGRESSION | ADHD comorbid with disruptive behaviour | (Demontis et al., 2021) | PGC | 3,802 cases + 31,305 controls |
| | Antisocial behaviour | (Tielbeek et al., 2017) | Broad ABC | 16,400 individuals |
| | Childhood aggression | (Ip et al., 2021) | Authors | 87,000 individuals |
| RELATED BEHAVIOURAL TRAITS | Anxiety | (Meier et al., 2019) | iPSYCH | 13,000 cases + 19,000 controls |
| | Anxiety: mental health problems ever diagnosed by a professional: Anxiety, nerves or generalized anxiety disorder | - | UKBiobank | 16,730 cases + 101,021 controls |
| | Anxiety: worrier / anxious feelings | - | UKBiobank | 199,463 cases + 152,370 controls |
| | "Ever been injured or injured someone else through drinking alcohol": Yes, during the last year | - | UKBiobank | 4,541 cases + 11,3461 controls |
| | Irritability | - | UKBiobank | 97,000 cases + 250,000 controls |
| | Neuroticism score | - | UKBiobank | 293,006 individuals |
| | Risk-taking behaviour | - | UKBiobank | 326,000 individuals |
| | Risk tolerance | (Karlsson Linnér et al., 2019) | Authors | 975,353 individuals |

ADHD, attention-deficit and hyperactivity disorder.

**Supplementary Table 3.** Heritability analyses using linkage disequilibrium score regression (LDSC).

| | **Trait or disorder** | **SNP heritability** | **Standard error** |
|---|---|---|---|
| **ADDICTION** | Alcohol dependence | 0.1098 | 0.0206 |
| | Cannabis dependence | **0.0308** | 0.0488 |
| | Cannabis use disorder | 0.061 | 0.0051 |
| | Cocaine dependence | 0.2661 | 0.0515 |
| | Ever addicted to any substance or behaviour | **0.0077** | 0.002 |
| | Opioids dependence | 0.1311 | 0.0514 |
| | Opioids use disorder | 0.238 | 0.0125 |
| | Substance use disorder | 0.139 | 0.0056 |
| **AGGRESSION** | ADHD comorbid with disruptive behaviour | 0.1461 | 0.0163 |
| | Antisocial behaviour | 0.0567 | 0.0263 |
| | Childhood aggression | **0.0371** | 0.0042 |
| **RELATED BEHAVIOURAL TRAITS** | Anxiety (iPSYCH) | **0.0192** | 0.0328 |
| | Anxiety: mental health problems ever diagnosed by a professional: Anxiety, nerves or generalized anxiety disorder | **0.0308** | 0.0036 |
| | Anxiety: worrier / anxious feelings | 0.098 | 0.005 |
| | "Ever been injured or injured someone else through drinking alcohol": "Yes, during the last year" | **0.0009** | 0.0012 |
| | Irritability | 0.0633 | 0.0034 |
| | Neuroticism score | 0.0884 | 0.0041 |
| | Risk-taking behaviour | 0.0549 | 0.0028 |
| | Risk tolerance | **0.0402** | 0.0015 |

ADHD, attention-deficit and hyperactivity disorder; In bold: SNP heritability lower than 5%.

**Supplementary Table 4**. Dopaminergic genes significantly associated with at least one phenotype in the gene-based analyses of 11 disorders or traits. All genes included in this table overcome a multiple-testing correction of FDR 5%.

| ADDICTION | | | | | | RELATED BEHAVIOURAL TRAITS | | | | | | | |
|---|---|---|---|---|---|---|---|---|---|---|---|---|---|
| Alcohol dependence | | OUD | | SUD | | Anxiety | | Irritability | | Neuroticism | | Risk-taking | |
| Gene name | p-value | Gene name | p-value | Gene name | p-value | Gene name | p-value | Gene name | p-value | Gene name | p-value | Gene name | p-value |
| ***ADH1C*** | 1.98E-07 | ***DRD2*** | 6.25E-08 | ***DRD2*** | 7.74E-13 | *ATF6B* | 4.62E-10 | *ARNTL* | 7.25E-07 | ***DRD2*** | 6.12E-13 | *AKT3* | 8.09E-08 |
| | | *PPP1CC* | 1.02E-06 | *ADH1C* | 2.20E-11 | *PPP2R3A* | 5.12E-06 | *HPRT1* | 2.05E-06 | *ARNTL* | 1.72E-09 | *GRIN2A* | 3.42E-05 |
| | | *ADH1C* | 5.36E-05 | *CHRNA4* | 2.46E-07 | *CACNA1C* | 1.52E-05 | ***DRD2*** | 3.41E-05 | *ATF6B* | 1.88E-09 | *ARNTL* | 1.55E-04 |
| | | *CNTNAP4* | 8.23E-05 | *GRIA1* | 4.97E-07 | *PRKCA* | 1.77E-05 | *TYR* | 4.63E-05 | *HPRT1* | 5.70E-08 | *MAPK8* | 2.54E-04 |
| | | *ADH4* | 3.08E-04 | *OTX2* | 7.46E-07 | ***DRD2*** | 3.10E-05 | *GPR37* | 1.86E-04 | *PRKCA* | 4.43E-07 | *GPR156* | 4.05E-04 |
| | | *OPRM1* | 6.47E-04 | *ADH1B* | 1.99E-06 | *ARNTL* | 6.99E-05 | *TAT* | 2.83E-04 | *KCNJ3* | 1.80E-06 | *ATF4* | 6.54E-04 |
| | | *CAMK2G* | 1.52E-03 | *CREB3L1* | 7.18E-06 | *PPP2R2B* | 9.68E-05 | *CELSR3* | 1.92E-03 | *CREB3L1* | 1.48E-05 | *GSK3B* | 7.36E-04 |
| | | | | *GNB5* | 1.85E-05 | *HTR1B* | 4.34E-04 | *TYRP1* | 2.31E-03 | *TYR* | 1.74E-05 | *PPP2R3C* | 1.11E-03 |
| | | | | *ADCY5* | 5.81E-05 | *MAPK10* | 6.82E-04 | *DLG4* | 2.39E-03 | *ATF4* | 2.08E-05 | | |
| | | | | *ADH5* | 6.08E-05 | *LRRK2* | 8.83E-04 | *GOT1* | 2.82E-03 | *PPP2R2B* | 2.80E-05 | | |
| | | | | *AKT3* | 1.79E-04 | *PPP2R5D* | 1.52E-03 | | | *NR4A2* | 5.95E-05 | | |
| | | | | *ADH4* | 2.56E-04 | *HPRT1* | 1.53E-03 | | | *OTX2* | 7.62E-05 | | |
| | | | | *CACNA1A* | 2.93E-04 | *PALM* | 1.81E-03 | | | *PPP2R3A* | 3.38E-04 | | |
| | | | | ***DBH*** | 3.20E-04 | *SNCA* | 1.89E-03 | | | *SNCA* | 5.54E-04 | | |
| | | | | *CALML4* | 1.35E-03 | *GABBR1* | 2.10E-03 | | | *ITPR3* | 6.17E-04 | | |
| | | | | *SCN1A* | 1.59E-03 | *ATP1A3* | 2.23E-03 | | | *MAPK10* | 7.84E-04 | | |
| | | | | *CHRNA6* | 2.23E-03 | *WNT5A* | 4.19E-03 | | | *ADH7* | 8.86E-04 | | |
| | | | | *CHRNB2* | 2.92E-03 | *LMX1B* | 4.68E-03 | | | *HTR1B* | 9.63E-04 | | |
| | | | | *GRIN2A* | 3.03E-03 | | | | | *GABBR1* | 1.98E-03 | | |
| | | | | *ADCY6* | 3.04E-03 | | | | | *GPR37* | 2.42E-03 | | |
| | | | | *LMX1B* | 3.23E-03 | | | | | *GNAO1* | 2.87E-03 | | |
| | | | | *ADH7* | 3.32E-03 | | | | | *GDNF* | 2.91E-03 | | |
| | | | | *HTR1B* | 3.43E-03 | | | | | ***DRD3*** | 3.38E-03 | | |
| | | | | *PPP3CA* | 3.60E-03 | | | | | *PALM* | 3.54E-03 | | |
| | | | | *CREB3* | 3.65E-03 | | | | | *SCN1A* | 4.20E-03 | | |
| | | | | *PTGS2* | 3.95E-03 | | | | | *MAPK9* | 4.31E-03 | | |
| | | | | *CREB3L4* | 4.57E-03 | | | | | *RSPO2* | 4.83E-03 | | |
| | | | | | | | | | | *CLOCK* | 5.39E-03 | | |
| | | | | | | | | | | *RGS8* | 6.13E-03 | | |
| | | | | | | | | | | *LRRK2* | 7.45E-03 | | |

In bold, genes belonging to the DA-core gene set. OUD, opioids use disorder; SUD, substance use disorder.

**Supplementary Table 5**. Serotoninergic genes associated with at least one phenotype in the gene-based analyses of 11 disorders or traits. All genes included in this table overcome a multiple-testing correction of FDR 5%.

| ADDICTION | | | | RELATED BEHAVIOURAL TRAITS | | | | | | | |
|---|---|---|---|---|---|---|---|---|---|---|---|
| OUD | | SUD | | Anxiety | | Irritability | | Neuroticism | | Risk-taking | |
| Gene name | p-value | Gene name | p-value | Gene name | p-value | Gene name | p-value | Gene name | p-value | Gene name | p-value |
| ***CHRNA3*** | 2.11E-04 | *CHRNA3* | 1.28E-15 | *GNAI2* | 2.56E-07 | ***HTR1E*** | 1.82E-04 | *PRKCA* | 4.43E-07 | *GRIN2A* | 3.42E-05 |
| | | *CYP2D6* | 5.17E-06 | *CACNA1C* | 1.52E-05 | *GPM6B* | 1.75E-03 | *GNAI2* | 1.23E-06 | *CHRNA3* | 1.06E-03 |
| | | *GNB5* | 1.85E-05 | *PRKCA* | 1.77E-05 | | | *KCNJ3* | 1.80E-06 | ***HTR4*** | 1.15E-03 |
| | | *ADCY5* | 5.81E-05 | ***HTR1B*** | 4.34E-04 | | | *NOS1* | 3.65E-06 | | |
| | | *KCND2* | 1.57E-04 | ***HTR6*** | 1.71E-03 | | | ***HTR1E*** | 5.89E-05 | | |
| | | *CACNA1A* | 2.93E-04 | *SNCA* | 1.89E-03 | | | *SNCA* | 5.54E-04 | | |
| | | *HADH* | 8.02E-04 | | | | | *ITPR3* | 6.17E-04 | | |
| | | ***HTR3A*** | 1.17E-03 | | | | | ***HTR1B*** | 9.63E-04 | | |
| | | ***HTR3B*** | 1.45E-03 | | | | | *KYNU* | 2.20E-03 | | |
| | | *GRIN2A* | 3.03E-03 | | | | | *MAPK3* | 2.75E-03 | | |
| | | *INMT* | 3.24E-03 | | | | | *GNAO1* | 2.87E-03 | | |
| | | ***HTR1B*** | 3.43E-03 | | | | | ***HTR4*** | 5.21E-03 | | |
| | | *PTGS2* | 3.95E-03 | | | | | | | | |

In bold, genes belonging to the 5-HT-core gene set. OUD, opioids use disorder; SUD, substance use disorder.

**Supplementary Table 6**. Results of the S-MultiXcan analysis.

| | DISORDER | Number of computed genes | Number of total significant genes (FDR 5%) | Number of DA significant genes (FDR 5%) | Number of 5-HT significant genes (FDR 5%) |
|---|---|---|---|---|---|
| AGGRESSION | AB | 14171 | 0 | 0 | 0 |
| | ADHD-DBD | 14164 | 0 | 0 | 0 |
| ADDICTION | CUD | 14193 | 2 | 0 | 0 |
| | OUD | 13069 | 212 | 7 | 3 |
| | SUD | 14175 | 339 | 10 | 4 |
| RELATED BEHAVIOURAL TRAITS | Anxiety: worrier / anxious feelings | 14201 | 309 | 5 | 1 |
| | Irritability | 14201 | 160 | 2 | 2 |
| | Neuroticism score | 14201 | 391 | 9 | 2 |
| | Risk-taking behaviour | 14200 | 48 | 0 | 0 |

5-HT, serotonin; AB, antisocial behaviour; ADHD-DBD, attention-deficit and hyperactivity disorder comorbid with disruptive behaviour; CUD, cannabis use disorder; DA, dopamine; OUD, opioids use disorder; SUD, substance use disorder; Risk-Taking; risk-taking behaviour. FDR, false discovery rate.

#### 2. SUPPLEMENTARY FIGURES

**ENIGMA**

| Gene sets | genes | VOLUME OF SUBCORTICAL AREAS | | | | | | | OTHER ANATOMICAL MEASURES | | |
|---|---|---|---|---|---|---|---|---|---|---|---|
| | | Amyg. | Caudate | Hipp. | NAcc. | Pallidum | Putamen | Thal. | ICV | Thick. | SA |
| DA core | 12 | 0 | 0 | 0 | 0 | 0 | 0 | 0 | 0 | 0 | **1** |
| DA wide | 275 | 0 | 0 | 0 | 0 | 0 | 0 | 0 | 0 | **1** | **3** |
| 5-HT core | 23 | 0 | 0 | 0 | 0 | 0 | 0 | 0 | 0 | 0 | **1** |
| 5-HT wide | 176 | 0 | 0 | 0 | 0 | 0 | 0 | 0 | 0 | 0 | **1** |
| All genes | | 0 | 0 | **2** | 0 | 0 | **6** | 0 | **11** | **21** | **103** |

**Supplementary Figure 1. Number of significant genes in the gene-based analyses of ENIGMA summary statistics of brain anatomical measures.** Significant genes have overcome a multiple-testing correction of FDR<5%. 5-HT, serotonin; DA, dopamine. Amyg., amygdala volume; Caudate, caudate volume; Hipp., hippocampus volume; ICV, intra-cranial volume; NAcc., nucleus accumbens volume; Pallidum, pallidum volume; Putamen, putamen volume; SA, surface area; Thal., thalamus volume; Thick., cortical thickness.

Ip, H.F., van der Laan, C.M., Krapohl, E.M.L., Brikell, I., Sánchez-Mora, C., Nolte, I.M., St Pourcain, B., Bolhuis, K., Palviainen, T., Zafarmand, H., Colodro-Conde, L., Gordon, S., Zayats, T., Aliev, F., Jiang, C., Wang, C.A., Saunders, G., Karhunen, V., Hammerschlag, A.R., Adkins, D.E., Border, R., Peterson, R.E., Prinz, J.A., Thiering, E., Seppälä, I., Vilor-Tejedor, N., Ahluwalia, T.S., Day, F.R., Hottenga, J.J., Allegrini, A.G., Rimfeld, K., Chen, Q., Lu, Y., Martin, J., Soler Artigas, M., Rovira, P., Bosch, R., Español, G., Ramos Quiroga, J.A., Neumann, A., Ensink, J., Grasby, K., Morosoli, J.J., Tong, X., Marrington, S., Middeldorp, C., Scott, J.G., Vinkhuyzen, A., Shabalin, A.A., Corley, R., Evans, L.M., Sugden, K., Alemany, S., Sass, L., Vinding, R., Ruth, K., Tyrrell, J., Davies, G.E., Ehli, E.A., Hagenbeek, F.A., De Zeeuw, E., Van Beijsterveldt, T.C.E.M., Larsson, H., Snieder, H., Verhulst, F.C., Amin, N., Whipp, A.M., Korhonen, T., Vuoksimaa, E., Rose, R.J., Uitterlinden, A.G., Heath, A.C., Madden, P., Haavik, J., Harris, J.R., Helgeland, Ø., Johansson, S., Knudsen, G.P.S., Njolstad, P.R., Lu, Q., Rodriguez, A., Henders, A.K., Mamun, A., Najman, J.M., Brown, S., Hopfer, C., Krauter, K., Reynolds, C., Smolen, A., Stallings, M., Wadsworth, S., Wall, T.L., Silberg, J.L., Miller, A., Keltikangas-Järvinen, L., Hakulinen, C., Pulkki-Råback, L., Havdahl, A., Magnus, P., Raitakari, O.T., Perry, J.R.B., Llop, S., Lopez-Espinosa, M.J., Bønnelykke, K., Bisgaard, H., Sunyer, J., Lehtimäki, T., Arseneault, L., Standl, M., Heinrich, J., Boden, J., Pearson, J., Horwood, L.J., Kennedy, M., Poulton, R., Eaves, L.J., Maes, H.H., Hewitt, J., Copeland, W.E., Costello, E.J., Williams, G.M., Wray, N., Järvelin, M.R., McGue, M., Iacono, W., Caspi, A., Moffitt, T.E., Whitehouse, A., Pennell, C.E., Klump, K.L., Burt, S.A., Dick, D.M., Reichborn-Kjennerud, T., Martin, N.G., Medland, S.E., Vrijkotte, T., Kaprio, J., Tiemeier, H., Davey Smith, G., Hartman, C.A., Oldehinkel, A.J., Casas, M., Ribasés, M., Lichtenstein, P., Lundström, S., Plomin, R., Bartels, M., Nivard, M.G., Boomsma, D.I., 2021. Genetic association study of childhood aggression across raters, instruments, and age. Transl. Psychiatry 11. https://doi.org/10.1038/s41398-021-01480-x

Johnson, E.C., Demontis, D., Thorgeirsson, T.E., Walters, R.K., Polimanti, R., Hatoum, A.S., Sanchez-Roige, S., Paul, S.E., Wendt, F.R., Clarke, T.K., Lai, D., Reginsson, G.W., Zhou, H., He, J., Baranger, D.A.A., Gudbjartsson, D.F., Wedow, R., Adkins, D.E., Adkins, A.E., Alexander, J., Bacanu, S.A., Bigdeli, T.B., Boden, J., Brown, S.A., Bucholz, K.K., Bybjerg-Grauholm, J., Corley, R.P., Degenhardt, L., Dick, D.M., Domingue, B.W., Fox, L., Goate, A.M., Gordon, S.D., Hack, L.M., Hancock, D.B., Hartz, S.M., Hickie, I.B., Hougaard, D.M., Krauter, K., Lind, P.A., McClintick, J.N., McQueen, M.B., Meyers, J.L., Montgomery, G.W., Mors, O., Mortensen, P.B., Nordentoft, M., Pearson, J.F., Peterson, R.E., Reynolds, M.D., Rice, J.P., Runarsdottir, V., Saccone, N.L., Sherva, R., Silberg, J.L., Tarter, R.E., Tyrfingsson, T., Wall, T.L., Webb, B.T., Werge, T., Wetherill, L., Wright, M.J., Zellers, S., Adams, M.J., Bierut, L.J., Boardman, J.D., Copeland, W.E., Farrer, L.A., Foroud, T.M., Gillespie, N.A., Grucza, R.A., Harris, K.M., Heath, A.C., Hesselbrock, V., Hewitt, J.K.,

Hopfer, C.J., Horwood, J., Iacono, W.G., Johnson, E.O., Kendler, K.S., Kennedy, M.A., Kranzler, H.R., Madden, P.A.F., Maes, H.H., Maher, B.S., Martin, N.G., McGue, Matthew, McIntosh, A.M., Medland, S.E., Nelson, E.C., Porjesz, B., Riley, B.P., Stallings, M.C., Vanyukov, M.M., Vrieze, S., Walters, R., Johnson, Emma, McClintick, J., Hatoum, A., Wendt, F., Adams, M., Adkins, A., Aliev, F., Batzler, A., Bertelsen, S., Biernacka, J., Bigdeli, T., Chen, L.S., Chou, Y.L., Degenhardt, F., Docherty, A., Edwards, A., Fontanillas, P., Foo, J., Frank, J., Giegling, I., Gordon, S., Hack, L., Hartmann, A., Heilmann-Heimbach, S., Herms, S., Hodgkinson, C., Hoffman, P., Hottenga, J., Kennedy, M., Alanne-Kinnunen, M., Konte, B., Lahti, J., Lahti-Pulkkinen, M., Ligthart, L., Loukola, A., Maher, B., Mbarek, H., McQueen, M., Meyers, J., Milaneschi, Y., Palviainen, T., Pearson, J., Peterson, R., Ripatti, S., Ryu, E., Saccone, N., Salvatore, J., Schwandt, M., Streit, F., Strohmaier, J., Thomas, N., Wang, J.C., Webb, B., Wills, A., Boardman, J., Chen, D., Choi, D.S., Copeland, W., Culverhouse, R., Dahmen, N., Domingue, B., Elson, S., Frye, M., Gäbel, W., Hayward, C., Ising, M., Keyes, M., Kiefer, F., Kramer, J., Kuperman, S., Lucae, S., Lynskey, M., Maier, W., Mann, K., Männistö, S., Müller-Myhsok, B., Murray, A., Nurnberger, J., Palotie, A., Preuss, U., Räikkönen, K., Reynolds, M., Ridinger, M., Scherbaum, N., Schuckit, M., Soyka, M., Treutlein, J., Witt, S., Wodarz, N., Zill, P., Adkins, D., Boomsma, D., Brown, S., Cichon, S., Costello, E.J., de Wit, H., Diazgranados, N., Dick, D., Eriksson, J., Farrer, L., Foroud, T., Gillespie, N., Goate, A., Goldman, D., Grucza, R., Hancock, D., Hewitt, J., Hopfer, C., Iacono, W., Johnson, Eric, Kaprio, J., Karpyak, V., Kranzler, H., Lichtenstein, P., Lind, P., McGue, Matt, MacKillop, J., Maes, H., Magnusson, P., Martin, N., Montgomery, G., Nelson, E., Nöthen, M., Palmer, A., Pederson, N., Penninx, B., Rice, J., Rietschel, M., Riley, B., Rose, R., Rujescu, D., Shen, P.H., Silberg, J., Tarter, R., Vanyukov, M., Wall, T., Whitfield, J., Zhao, H., Neale, B., Gelernter, J., Edenberg, H., Agrawal, A., Davis, L.K., Bogdan, R., Edenberg, H.J., Stefansson, K., Børglum, A.D., 2020. A large-scale genome-wide association study meta-analysis of cannabis use disorder. The Lancet Psychiatry 7, 1032–1045. https://doi.org/10.1016/S2215-0366(20)30339-4

Karlsson Linnér, R., Biroli, P., Kong, E., Meddens, S.F.W., Wedow, R., Fontana, M.A., Lebreton, M., Tino, S.P., Abdellaoui, A., Hammerschlag, A.R., Nivard, M.G., Okbay, A., Rietveld, C.A., Timshel, P.N., Trzaskowski, M., Vlaming, R. de, Zünd, C.L., Bao, Y., Buzdugan, L., Caplin, A.H., Chen, C.Y., Eibich, P., Fontanillas, P., Gonzalez, J.R., Joshi, P.K., Karhunen, V., Kleinman, A., Levin, R.Z., Lill, C.M., Meddens, G.A., Muntané, G., Sanchez-Roige, S., Rooij, F.J. va., Taskesen, E., Wu, Y., Zhang, F., Agee, M., Alipanahi, B., Bell, R.K., Bryc, K., Elson, S.L., Furlotte, N.A., Huber, K.E., Litterman, N.K., McCreight, J.C., McIntyre, M.H., Mountain, J.L., Northover, C.A.M., Pitts, S.J., Sathirapongsasuti, J.F., Sazonova, O. V., Shelton, J.F., Shringarpure, S., Tian, C., Tung, J.Y., Vacic, V., Wilson, C.H., Agbessi, M., Ahsan, H., Alves, I., Andiappan, A., Awadalla, P., Battle, A., Beutner, F., Jan Bonder, M., Boomsma, D.I., Christiansen, M., Claringbould, A., Deelen, P., Esko, T., Favé, M.J., Franke, L., Frayling, T., Gharib, S.A., Gibson, G., Heijmans, B., Hemani, G., Jansen, R., Kähönen, M., Kalnapenkis, A., Kasela, S., Kettunen, J., Kim, Y., Kirsten, H., Kovacs, P., Krohn, K., Kronberg-Guzman, J., Kukushkina, V., Kutalik, Z., Lee, B., Lehtimäki, T., Loeffler, M., Marigorta, U.M., Metspalu, A., Milani, L., Montgomery, G.W., Müller-Nurasyid, M., Nauck, M., Penninx, B., Perola, M., Pervjakova, N., Pierce, B., Powell, J., Prokisch, H., Psaty, B.M., Raitakari, O., Ring, S., Ripatti, S., Rotzchke, O., Rüeger, S., Saha, A., Scholz, M., Schramm, K., Seppälä, I., Stumvoll, M., Sullivan, P., Hoen, P.B. t., Teumer, A., Thiery, J., Tong, L., Tönjes, A., Dongen, J. van, Meurs, J. van, Verlouw, J., Visscher, P.M., Völker, U., Võsa, U., Westra, H.J., Yaghootkar, H., Yang, Jian, Zeng, B., Beauchamp, J.P., Lee, J.J., Pers, T.H., Turley, P., Chen, G.B., Emilsson, V., Oskarsson, S., Pickrell, J.K., Thom, K., Timshel, P., de Vlaming, R., Ahluwalia, T.S., Bacelis, J., Baumbach, C., Bjornsdottir, G., Brandsma, J.H., Concas, M.P., Derringer, J., Galesloot, T.E., Girotto, G., Gupta, R., Hall, L.M., Harris, S.E., Hofer, E., Horikoshi, M., Huffman, J.E., Kaasik, K., Kalafati, I.P., Karlsson, R., Kong, A., Lahti, J., Lee, S.J. va. der, de Leeuw, C., Lind, P.A., Lindgren, K.O., Liu, T., Mangino, M., Marten, J., Mihailov, E., Miller, M.B., Most, P.J. va. der, Oldmeadow, C., Payton, A., Peyrot, W.J., Qian, Y., Rueedi, R., Salvi, E., Schmidt, B., Schraut, K.E., Shi, J., Smith, A. V., Poot, R.A., Pourcain, B.S., Thorleifsson, G., Verweij, N., Vuckovic, D., Wellmann, J., Yang, Jingyun, Zhao, W., Zhu, Z., Alizadeh, B.Z., Amin, N., Bakshi, A., Baumeister, S.E., Biino, G., Bønnelykke, K., Boyle, P.A., Campbell, H., Cappuccio, F.P., Davies, G., De Neve, J.E., Deloukas, P., Demuth, I., Ding, J., Eisele, L., Eklund, N., Evans, D.M., Faul, J.D., Feitosa, M.F., Forstner, A.J., Gandin, I., Gunnarsson, B., Halldórsson, B. V., Harris, T.B., Heath, A.C., Hocking, L.J., Holliday, E.G., Homuth, G., Horan, M.A., Hottenga, J.J., de Jager, P.L., Jugessur, A., Kaakinen, M.A., Kanoni, S., Keltigangas-Järvinen, L., Kiemeney, L.A.L.M., Kolcic, I., Koskinen, S., Kraja, A.T., Kroh, M., Latvala, A., Launer, L.J., Lebreton, M.P., Levinson, D.F., Lichtenstein, P., Lichtner, P., Liewald, D.C.M., Loukola, A., Madden, P.A., Mägi, R., Mäki-Opas, T., Marioni, R.E., Marques-Vidal, P., McMahon, G., Meisinger, C., Meitinger, T., Milaneschi, Y., Myhre, R., Nelson, C.P., Nyholt, D.R., Ollier, W.E.R., Palotie, A., Paternoster, L., Pedersen, N.L., Petrovic, K.E., Porteous, D.J., Räikkönen, K., Ring, S.M., Robino, A., Rostapshova, O.,

Rudan, I., Rustichini, A., Salomaa, V., Sanders, A.R., Sarin, A.P., Schmidt, H., Scott, R.J., Smith, B.H., Smith, J.A., Staessen, J.A., Steinhagen-Thiessen, E., Strauch, K., Terracciano, A., Tobin, M.D., Ulivi, S., Vaccargiu, S., Quaye, L., Venturini, C., Vinkhuyzen, A.A.E., Völzke, H., Vonk, J.M., Vozzi, D., Waage, J., Ware, E.B., Willemsen, G., Attia, J.R., Bennett, D.A., Berger, K., Bertram, L., Bisgaard, H., Borecki, I.B., Bültmann, U., Chabris, C.F., Cucca, F., Cusi, D., Deary, I.J., Dedoussis, G. V., Duijn, C.M. va., Eriksson, J.G., Franke, B., Gasparini, P., Gejman, P. V., Gieger, C., Grabe, H.J., Gratten, J., Groenen, P.J.F., Gudnason, V., Harst, P. van der, Hayward, C., Hinds, D.A., Hoffmann, W., Hyppönen, E., Iacono, W.G., Jacobsson, B., Järvelin, M.R., Jöckel, K.H., Kaprio, J., Kardia, S.L.R., Lehrer, S.F., Magnusson, P.K.E., Martin, N.G., McGue, M., Pendleton, N., Pirastu, N., Pirastu, M., Polasek, O., Posthuma, D., Power, C., Province, M.A., Samani, N.J., Schlessinger, D., Schmidt, R., Sørensen, T.I.A., Spector, T.D., Stefansson, K., Thorsteinsdottir, U., Thurik, A.R., Timpson, N.J., Tiemeier, H., Uitterlinden, A.G., Vitart, V., Vollenweider, P., Weir, D.R., Wilson, J.F., Wright, A.F., Conley, D.C., Krueger, R.F., Smith, G.D., Hofman, A., Laibson, D.I., Medland, S.E., Meyer, M.N., Johannesson, M., Koellinger, P.D., Cesarini, D., Benjamin, D.J., Auton, A., Boardman, J.D., Clark, D.W., Conlin, A., Dolan, C.C., Fischbacher, U., Harris, K.M., Hasler, G., Ikram, M.A., Jain, S., Kessler, R.C., Kooyman, M., MacKillop, J., Männikkö, M., Morcillo-Suarez, C., McQueen, M.B., Schmidt, K.M., Smart, M.C., Sutter, M., White, J., Wit, H. de, Fehr, E., Kumari, M., Navarro, A., Palmer, A.A., Schunk, D., Stein, M.B., Svento, R., Timmers, P.R.H.J., Ursano, R.J., Wagner, G.G., 2019. Genome-wide association analyses of risk tolerance and risky behaviors in over 1 million individuals identify hundreds of loci and shared genetic influences. Nat. Genet. 51, 245–257. https://doi.org/10.1038/s41588-018-0309-3

Meier, S.M., Trontti, K., Purves, K.L., Als, T.D., Grove, J., Laine, M., Pedersen, M.G., Bybjerg-Grauholm, J., Bækved-Hansen, M., Sokolowska, E., Mortensen, P.B., Hougaard, D.M., Werge, T., Nordentoft, M., Breen, G., Børglum, A.D., Eley, T.C., Hovatta, I., Mattheisen, M., Mors, O., 2019. Genetic Variants Associated with Anxiety and Stress-Related Disorders: A Genome-Wide Association Study and Mouse-Model Study. JAMA Psychiatry 76, 924–932. https://doi.org/10.1001/jamapsychiatry.2019.1119

Polimanti, R., Walters, R.K., Johnson, E.C., McClintick, J.N., Adkins, A.E., Adkins, D.E., Bacanu, S.A., Bierut, L.J., Bigdeli, T.B., Brown, S., Bucholz, K.K., Copeland, W.E., Costello, E.J., Degenhardt, L., Farrer, L.A., Foroud, T.M., Fox, L., Goate, A.M., Grucza, R., Hack, L.M., Hancock, D.B., Hartz, S.M., Heath, A.C., Hewitt, J.K., Hopfer, C.J., Johnson, E.O., Kendler, K.S., Kranzler, H.R., Krauter, K., Lai, D., Madden, P.A.F., Martin, N.G., Maes, H.H., Nelson, E.C., Peterson, R.E., Porjesz, B., Riley, B.P., Saccone, N., Stallings, M., Wall, T.L., Webb, B.T., Wetherill, L., Edenberg, H.J., Agrawal, A., Gelernter, J., 2020. Leveraging genome-wide data to investigate differences between opioid use vs. opioid dependence in 41,176 individuals from the Psychiatric Genomics Consortium. Mol. Psychiatry 25, 1673–1687. https://doi.org/10.1038/s41380-020-0677-9

Schoeler, T., Baldwin, J., Allegrini, A., Barkhuizen, W., McQuillin, A., Pirastu, N., Kutalik, Z., Pingault, J.B., 2022. Novel Biological Insights Into the Common Heritable Liability to Substance Involvement: A Multivariate Genome-wide Association Study. Biol. Psychiatry 93, 524–535. https://doi.org/10.1016/j.biopsych.2022.07.027

Sherva, R., Wang, Q., Kranzler, H., Zhao, H., Koesterer, R., Herman, A., Farrer, L.A., Gelernter, J., 2016. Genome-wide association study of cannabis dependence severity, novel risk variants, and shared genetic risks. JAMA Psychiatry 73, 472–480. https://doi.org/10.1001/jamapsychiatry.2016.0036

Tielbeek, J.J., Johansson, A., Polderman, T.J.C., Rautiainen, M.R., Jansen, P., Taylor, M., Tong, X., Lu, Q., Burt, A.S., Tiemeier, H., Viding, E., Plomin, R., Martin, N.G., Heath, A.C., Madden, P.A.F., Montgomery, G., Beaver, K.M., Waldman, I., Gelernter, J., Kranzler, H.R., Farrer, L.A., Perry, J.R.B., Munafò, M., LoParo, D., Paunio, T., Tiihonen, J., Mous, S.E., Pappa, I., De Leeuw, C., Watanabe, K., Hammerschlag, A.R., Salvatore, J.E., Aliev, F., Bigdeli, T.B., Dick, D., Faraone, S. V., Popma, A., Medland, S.E., Posthuma, D., 2017. Genome-wide association studies of a broad spectrum of antisocial behavior. JAMA Psychiatry 74, 1242–1250. https://doi.org/10.1001/jamapsychiatry.2017.3069

Walters, R.K., Polimanti, R., Johnson, E.C., McClintick, J.N., Adams, M.J., Adkins, A.E., Aliev, F., Bacanu, S.A., Batzler, A., Bertelsen, S., Biernacka, J.M., Bigdeli, T.B., Chen, L.S., Clarke, T.K., Chou, Y.L., Degenhardt, F., Docherty, A.R., Edwards, A.C., Fontanillas, P., Foo, J.C., Fox, L., Frank, J., Giegling, I., Gordon, S., Hack, L.M., Hartmann, A.M., Hartz, S.M., Heilmann-Heimbach, S., Herms, S., Hodgkinson, C., Hoffmann, P., Jan Hottenga, J., Kennedy, M.A., Alanne-Kinnunen, M., Konte, B., Lahti, J., Lahti-Pulkkinen, M., Lai, D., Ligthart, L., Loukola, A., Maher, B.S., Mbarek, H., McIntosh, A.M., McQueen, M.B., Meyers, J.L.,

Milaneschi, Y., Palviainen, T., Pearson, J.F., Peterson, R.E., Ripatti, S., Ryu, E., Saccone, N.L., Salvatore, J.E., Sanchez-Roige, S., Schwandt, M., Sherva, R., Streit, F., Strohmaier, J., Thomas, N., Wang, J.C., Webb, B.T., Wedow, R., Wetherill, L., Wills, A.G., Agee, M., Alipanahi, B., Auton, A., Bell, R.K., Bryc, K., Elson, S.L., Fontanillas, P., Furlotte, N.A., Hinds, D.A., Huber, K.E., Kleinman, A., Litterman, N.K., McCreight, J.C., McIntyre, M.H., Mountain, J.L., Noblin, E.S., Northover, C.A.M., Pitts, S.J., Sathirapongsasuti, J.F., Sazonova, O. V., Shelton, J.F., Shringarpure, S., Tian, C., Tung, J.Y., Vacic, V., Wilson, C.H., Boardman, J.D., Chen, D., Choi, D.S., Copeland, W.E., Culverhouse, R.C., Dahmen, N., Degenhardt, L., Domingue, B.W., Elson, S.L., Frye, M.A., Gäbel, W., Hayward, C., Ising, M., Keyes, M., Kiefer, F., Kramer, J., Kuperman, S., Lucae, S., Lynskey, M.T., Maier, W., Mann, K., Männistö, S., Müller-Myhsok, B., Murray, A.D., Nurnberger, J.I., Palotie, A., Preuss, U., Räikkönen, K., Reynolds, M.D., Ridinger, M., Scherbaum, N., Schuckit, M.A., Soyka, M., Treutlein, J., Witt, S., Wodarz, N., Zill, P., Adkins, D.E., Boden, J.M., Boomsma, D.I., Bierut, L.J., Brown, S.A., Bucholz, K.K., Cichon, S., Costello, E.J., de Wit, H., Diazgranados, N., Dick, D.M., Eriksson, J.G., Farrer, L.A., Foroud, T.M., Gillespie, N.A., Goate, A.M., Goldman, D., Grucza, R.A., Hancock, D.B., Harris, K.M., Heath, A.C., Hesselbrock, V., Hewitt, J.K., Hopfer, C.J., Horwood, J., Iacono, W., Johnson, E.O., Kaprio, J.A., Karpyak, V.M., Kendler, K.S., Kranzler, H.R., Krauter, K., Lichtenstein, P., Lind, P.A., McGue, M., MacKillop, J., Madden, P.A.F., Maes, H.H., Magnusson, P., Martin, N.G., Medland, S.E., Montgomery, G.W., Nelson, E.C., Nöthen, M.M., Palmer, A.A., Pedersen, N.L., Penninx, B.W.J.H., Porjesz, B., Rice, J.P., Rietschel, M., Riley, B.P., Rose, R., Rujescu, D., Shen, P.H., Silberg, J., Stallings, M.C., Tarter, R.E., Vanyukov, M.M., Vrieze, S., Wall, T.L., Whitfield, J.B., Zhao, H., Neale, B.M., Gelernter, J., Edenberg, H.J., Agrawal, A., 2018. Transancestral GWAS of alcohol dependence reveals common genetic underpinnings with psychiatric disorders. Nat. Neurosci. 21, 1656–1669. https://doi.org/10.1038/s41593-018-0275-1
